# Clinical validation of pneumatic transportation systems for monoclonal antibodies

**DOI:** 10.1101/2023.03.25.23287739

**Authors:** Pierre Coliat, Stéphane Erb, Hélène Diemer, Dan Karouby, Mainak Banerjee, Chen Zhu, Martin Demarchi, Sarah Cianférani, Alexandre Detappe, Xavier Pivot

## Abstract

Pneumatic transportation systems (PTS) were recently proposed as a method to carry ready-for-injection diluted monoclonal antibodies (mAbs) from the pharmacy to the bedside of patients. This method reduces transportation time and improves the efficiency of drug distribution process. However, mAbs are highly sensitive molecules for which subtle alterations may lead to deleterious clinical effects. These alterations can be caused by various external factors such as temperature, pH, pressure, and mechanical forces that may occur during transportation. Hence, it is essential to ensure that the mAbs transported by PTS remain stable and active throughout the transportation process. This study aims to determine the safety profile of PTS to transport 11 routinely used mAbs in a clinical setting through assessment of critical quality attributes (CQA) and orthogonal analysis. Hence, we performed aggregation/degradation profiling, post-translational modifications identification using complementary mass spectrometry-based methods, along with visible and subvisible particle formation determination by light absorbance and dynamic light scattering measurements. Altogether, these results highlight that PTS can be safely used for this purpose when air is removed from the bags during preparation.

## Introduction

Over the past two decades, numerous therapeutic breakthroughs in oncology have been achieved with monoclonal antibodies (mAbs). These mAbs were developed and approved either as modulators of the oncogenic signaling pathway by targeting overexpressed receptors (*e.g*. trastuzumab and pertuzumab for ErbB2, cetuximab and panitumumab for ErbB1), as a trap of soluble ligands involved in angiogenesis (bevacizumab for VEGFA), or as immunomodulators by inhibiting immune checkpoints (pembrolizumab, atezolizumab, nivolumab, avelumab, and durvalumab for the PD-1/PD-L1 pathway, ipilimumab for the CTLA-4 pathway).

Unlike conventional small molecule chemotherapies, the intricate structures of mAbs make them susceptible to chemical modifications under conditions of chemical or physical stress^1^. Among chemical instabilities, methionine, histidine, and cysteine are known to cause changes in protein structure, potentially leading to oxidation and the formation of disulfide bonds^2^. Presence of deamidation of glutamine and asparagine can induce changes in the structure of the protein; as a result, the fragmentation of disulfide bonds may dissociate the mAbs structure and lead to an inactive product^3^.

The physical instabilities are mainly characterized by reversible and nonreversible aggregations due to weak nonspecific bond formation^1^. For those reasons, manufacturers need to perform extensive stability studies on the final product as well as on its diluted version which is specifically prepared for injection in a clinical setting to attain authorization for routine usage from regulatory agencies; these guidelines are well established and comparable worldwide^4,5^. Diluted mAbs products may undergo changing conditions; the reconstituted product contains lower concentrations of stabilizers (*i.e*., polysorbate 20, polysorbate 80) and additional excipients in the solvent (*i.e*., sodium chloride or dextrose). Thus, the manufacturers recommend that the product is administered within 24h after the dilution to minimize the risk of mAbs being altered.; In this study, the container is a plastic bag instead of a glass vial which exhibits a wider air-liquid interface that may induces aggregates and the formation of visible/subvisible particles due to the shaking of the container during transportation^6^. In addition, several studies have demonstrated that the lack of PS20 or PS80 increases the adhesion and aggregation of mAbs to the plastic bag in polyvinylchloride (PVC) or polyolefin^7–9^. These 2 major mAbs modifications are considered risks, as they may lead to immunogenicity^10^. The key critical quality attributes (CQAs) of mAbs, including the protein structure or the glycosylation profile, can be altered but cause a risk of altered activity and/or toxicity^11,12^; thus the conditions of preparation for mAbs must be considered with caution to ensure that the administrated therapy is safe and effective.

One of the most common feature present in all major hospitals is the presence of pneumatic transportation systems (PTS). They were originally developed to accelerate the transportation biological samples from one floor to another. More recently, they were deployed for the transportation of anticancer therapies. According to each country, the regulations supporting the routine use of PTS vary from a definitive prohibition guided by precautionary principles due to the lack of robust evidence to a systematic utilization. These variable approaches and discrepancies in the routine use of PTS occur because of the external stress applied to these mAbs during transportation as they undergo acceleration and deceleration of multiple g forces as well as radical gravity forces. Here, we sought to determine the possible alterations that occur in terms of CQA to qualify the routine use of PTS with 11 mAbs approved for clinical use in oncology by the European Medicines Agency (EMA).

## Results

### Characteristics of the PTS

The PTS from the Institut de Cancérologie Strasbourg Europe (ICANS) is an Aerocom AC4000 160mm millimeters, 1000m long and includes approx. 20 curves (**Figure 1A-B**). The cartridge used to transport the mAbs along the different departments from the institution undergoes an acceleration profile of 4.06 ± 1.75g *per* travel with more than 15 acceleration peaks of >20g *per* travel and a maximum at 32g that is reached 4 times (**Figure 1C**). For this study, we placed the diluted mAbs inside the cartridge; the mAbs went through the whole tube 1x to characterize the institution setup and 10x to generalize the results to almost any PTS system installed in Europe using the same technical features.

**Figure 1.**
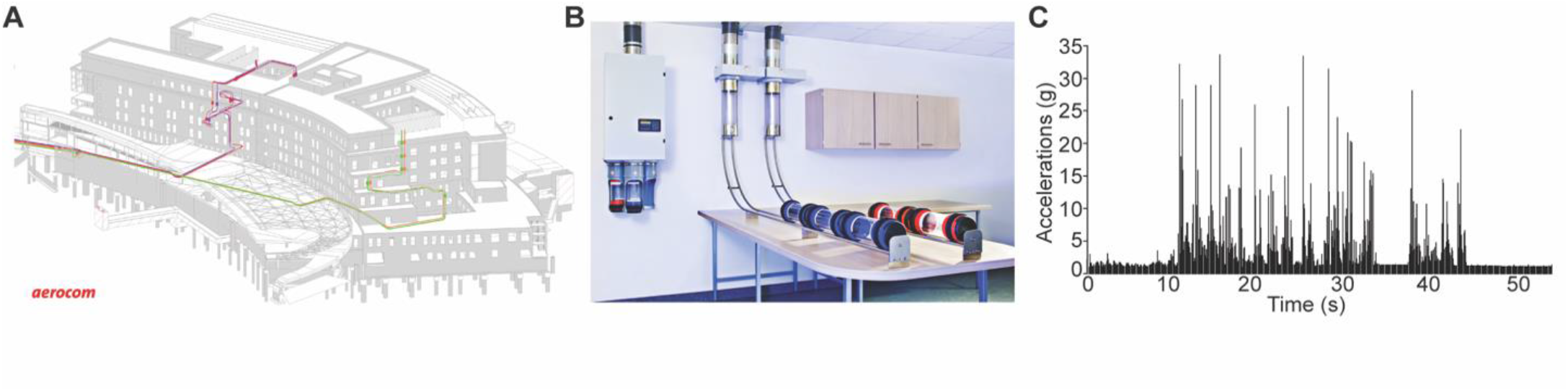
Characterization of the pneumatic transportation system (PTS). **A**. Map of the PTS at the Institut de Cancérologie Strasbourg Europe. In red the one used for this study. **B**. Representation of the setup used for transportation of the monoclonal antibodies (mAbs). **C**. Characterization of the cumulative (x, y, and z axis) acceleration forces applied to the mAbs.

### Aggregation analysis

We assessed the potential aggregation by characterizing the high molecular weight species (HMWS) and the degradation products by assessing the low molecular weight species (LMWS) by size-exclusion chromatography coupled to native mass spectrometry (SEC-nMS), a method that accurately quantifies HMWS/LMWS from SEC along with mass identification of each species within the same analysis^13,14^. The SEC-nMS was used to evaluate the HMWS and LMWS contained in the reference mAbs (non-diluted, in glass vial), post thermal stress (20 days 40°C, non-diluted, in glass vial, see material and methods), and for the diluted product passed 1-time in the PTS. We highlighted the results obtained with the mAb pertuzumab; these results will be used as case study throughout the manuscript, additional mAbs are presented as bar graphs. The SEC-nMS reveals 3 peaks corresponding to the main monomeric drug product (peak 2, MW 148,095 ± 2 Da, 99.8%), minor dimeric HMWS (peak 1, < 0.1%) and Fc-Fab LMWS (peak 3, <0.1 %). Samples after 1 PTS passage exhibit superimposable SEC-UV and nMS profiles to the one of reference samples, while thermally stressed samples show significantly higher amounts of both HMWS (dimers, 0.9%, p=0.027) and LMWS (100,941 ± 7 Da for Fc-Fab and 47,650 ± 2 Da for Fab fragments, 3.5% in total; p=0.0104) (**Figure 2A-B**).

**Figure 2:**
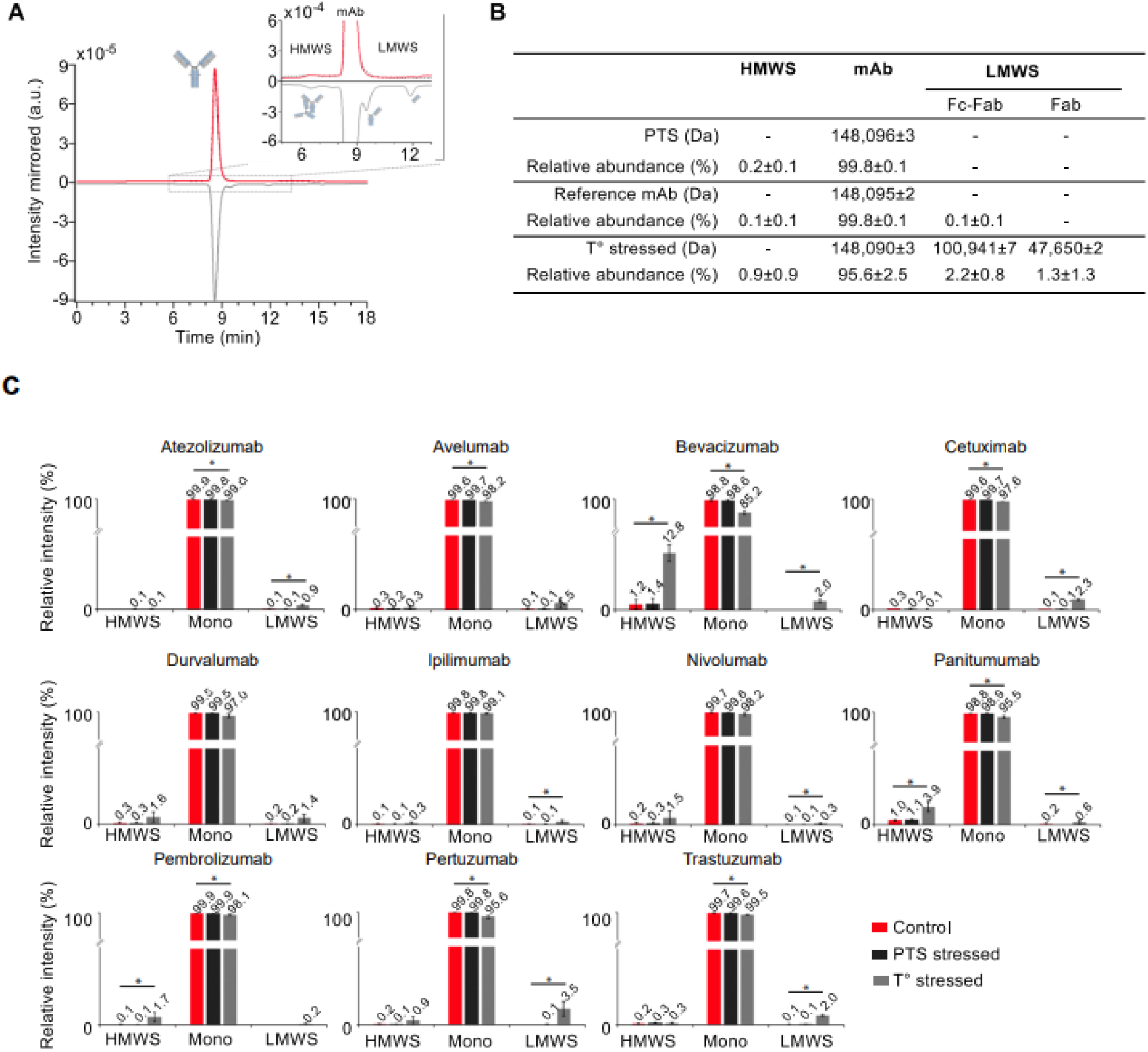
Intact level SEC-nMS analysis. **A**. Intact level SEC-nMS analysis of pertuzumab. SEC-UV chromatograms. Insert highlight the low levels of HMWS and LMWS. **B**. Summary of the masses measured for the main drug product and aggregates/fragments in the different conditions. **C**. Quantification of HMWS and LMWS from intact SEC-native MS analysis. Histograms represent SEC-UV peak area integration of aggregates (HMWS, mostly dimers) and fragments (mostly Fab-FC and Fab) for the 11 mAbs under investigation. *<u>Red :</u> Reference mAbs* ; *<u>Black</u>* : *1 pass-PTS mAbs, <u>Grey :</u> Thermally stressed mAbs*.

For all mAbs, the main observed peak was attributed to the monomeric mAb drug product highlighting high levels of purity for reference mAbs (> 99.5%, **Figure 2C**). Only very minor species corresponding to the mAb aggregates (dimers ∼300 kDa, < 0.3%) and fragments (Fab fragment ∼47 kDa, < 0.1%) were quantified from SEC-UV data and online identified by nMS. Only two mAbs (panitumumab and bevacizumab) showed a higher content of HMWS (∼1.1-1.2%) (**Figure 2C**), which is in agreement with literature data^15^.

After 1 PTS passage, chromatographic profiles were not significantly different from those of reference samples, and the drug substance being detected as main product (> 99.5%) with minor HMWS (<0.3%) and LMWS (<0.1%). The overall degradation profile was strongly affected upon thermal stress (higher HWMS and/or LMWS) independently of the 11 mAb compared to PTS and reference samples, confirming that the thermal stress should be used as positive control for mAb degradation^16^. Of interest, panitumumab, bevacizumab, trastuzumab, cetuximab and pertuzumab are significantly less stable under thermal stress, leading to either higher amounts of HMWS (> 3%, panitumumab and bevacizumab) or LMWS (>2%, trastuzumab, pertuzumab, bevacizumab and cetuximab).

Altogether, this study was performed with 11 diluted mAbs and demonstrated that neither reference mAbs, nor samples were degraded with one PTS passage in contrast to thermally stressed samples. They all exhibit comparable amounts of monomeric drug product (main peak), HMWS, and LMWS species confirming the safety profile of the PTS system from the ICANS institution. Similar results were obtained after 10 passages in the PTS for trastuzumab (**Figure S1**) confirming its safety profile.

### Glycoprofiles and post translational modifications (PTMs)

We next focused on glycoprofiles and some important PTMs which were defined as CQA (oxidations, N-terminal pyroglutamylation and C-terminal lysine deletion) to further examine the impact due to PTS at the molecular level. We combined complementary LC-MS methods in either native (SEC-nMS) or denaturing (reversed-phase liquid chromatography, rpLC-MS) conditions at both intact mAb and subunit levels (after IdeS digestion) to avoid extensive time-consuming peptide mapping (**Figure 3**).

**Figure 3:**
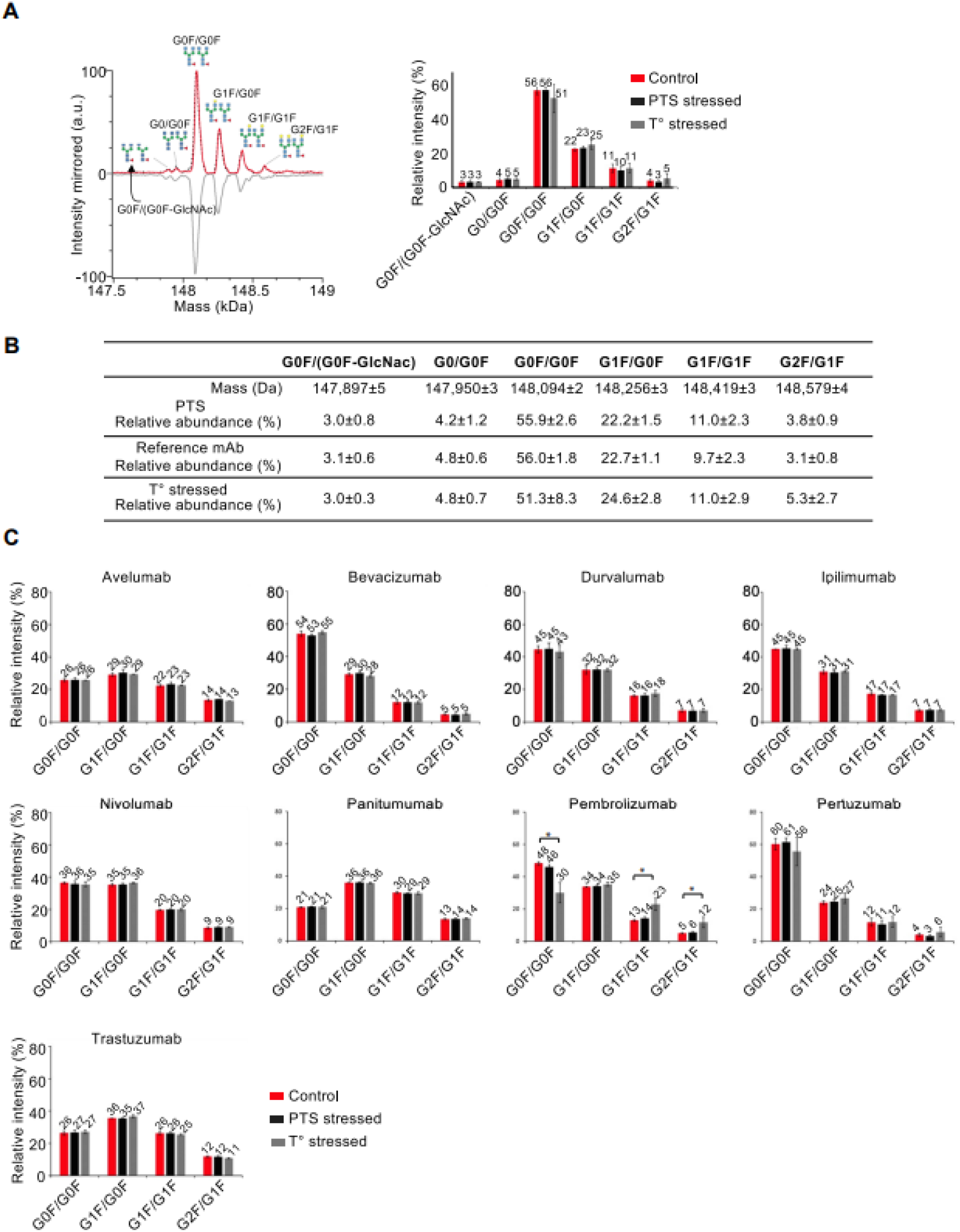
Glycoprofiles obtained from intact SEC-native MS analysis. **A**. Deconvoluted mass spectrum of mAb drug products (main peak) revealing glycoprofiles and corresponding histogram for glycoform quantification. **B**. Summary of the masses measured for main glycoforms. **C**. Histograms represent MS peak intensities obtained after zero-charge deconvolution of the main glycoforms for the 11 mAbs under investigation. Of note atezolizumab is an a-glycosylated mAb and not represented, while cetuximab glycoprofile could not be resolved using intact SEC-nMS. *<u>Red :</u> Reference mAbs* ; *<u>Black :</u> 1 pass-PTS mAbs, <u>Grey :</u> Thermally stressed mAbs*.

Glycosylation is among the most common PTM that can induce tremendous effects on the physicochemical properties, structure, and function of mAbs therapeutic efficiency^17^. The glycoprofile can affect the absorption distribution metabolization elimination (ADME) properties and the biosimilarity of the glycoprotein-based drugs by altering the PK profile^18^, receptor binding, and antibody dependent cell cytotoxicity (ADCC)^19^. Currently, MS-based techniques are widely used for the analysis of mAb heterogeneity of glycosylation^20,21^.

Glycoprofiles were first investigated using SEC-nMS on intact reference mAbs, after one PTS passage and after thermal stress. The overlaid deconvoluted native mass spectra of the 3 different pertuzumab samples (diluted, PTS, thermally stressed) demonstrates perfectly superimposable glycoprofiles at the intact level and unambiguous mass identification of the main glycovariants (G0F/G0F, G1F/G0F, G1F/G1F and G2F/G1F) (**Figure 3A-B**). No peak broadening is observed for the PTS samples in comparison to reference mAb conversely to thermally stressed samples. Among the 10 mAbs studied (atezolizumab being a-glycosylated), only cetuximab glycoprofiling could not be tackled by SEC-nMS, in agreement with this mAb bearing two glycosylation sites^22,23^. No significant glycoprofiles differences were observed for any of the samples in the different conditions (reference, PTS or thermally-stressed; p>0.05) (**Figure 3C**).

To cross validate the SEC-nMS results, the middle-up analysis of all mAbs were performed after IdeS digestion using a more classical high resolution rpLC-MS method on ∼25 kDa subunits^24^. Three main peaks are observed for pertuzumab, corresponding to Fc/2 (peak 2, 25236 Da), LC (peak 3, 23526), and Fd (peak 3, 25315) subunits respectively. The Fc/2 part bearing the N-glycosylation pattern (Man5, G0F-GlcNAc, G0, G0F, G1F, G2F) (**Figure 4**). The rpLC chromatogram of reference and PTS samples present similar patterns, whereas the peak 1 is more intense in the thermally stressed sample. Similar to the intact mass analysis, Fc/2 glycoprofiles of reference versus PTS or temperature stressed samples overlay perfectly.

**Figure 4:**
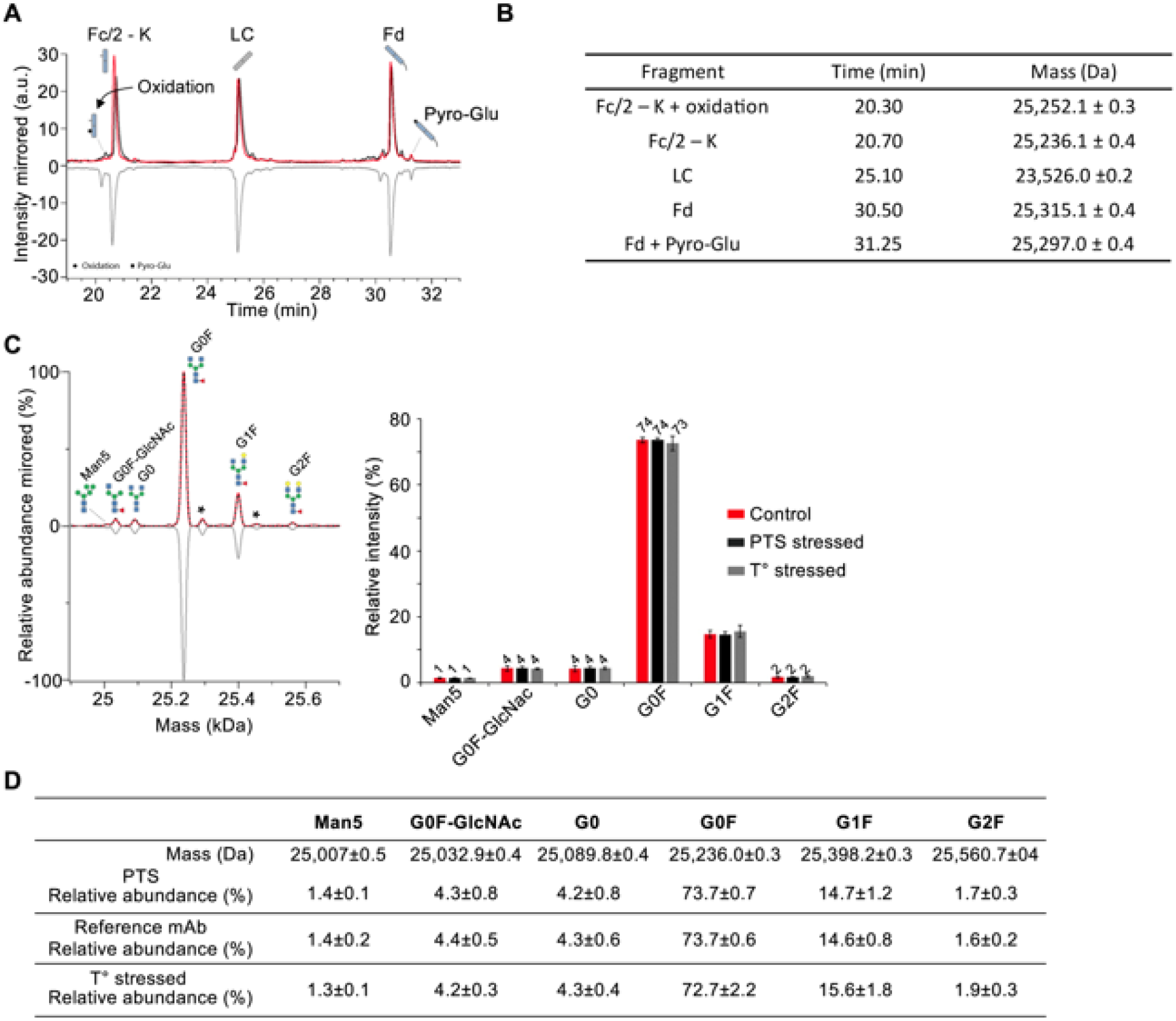
Middle level rpLC-MS analysis of pertuzumab. **A**. rpLC chromatogram of pertuzumab obtained after IdeS digestion highlighting the LC, Fd and Fc/2 subunits. **B**. Summary of the masses measured for the different subunits. **C**. Deconvoluted mass spectrum and of the Fc subunit revealing similar glycoprofiles and subsequent glycoform quatification presented as histograms. **D**. Summary of the masses of the main glycoforms. *<u>Red :</u> Reference mAbs* ; *<u>Black :</u> 1 pass-PTS mAbs, <u>Grey :</u> Thermally stressed mAbs*.

The glycoprofiles observed for all 10 other mAbs were non-significantly different (**Figure 5**), suggesting that the glycoprofiles are not significantly affected upon PTS passage (p>0.05). Interestingly, bevacizumab undergoes loss of G0F upon thermal stress. This result is in accordance with knowledge that this mAb is more prone to stress conditions than other mAbs.

**Figure 5:**
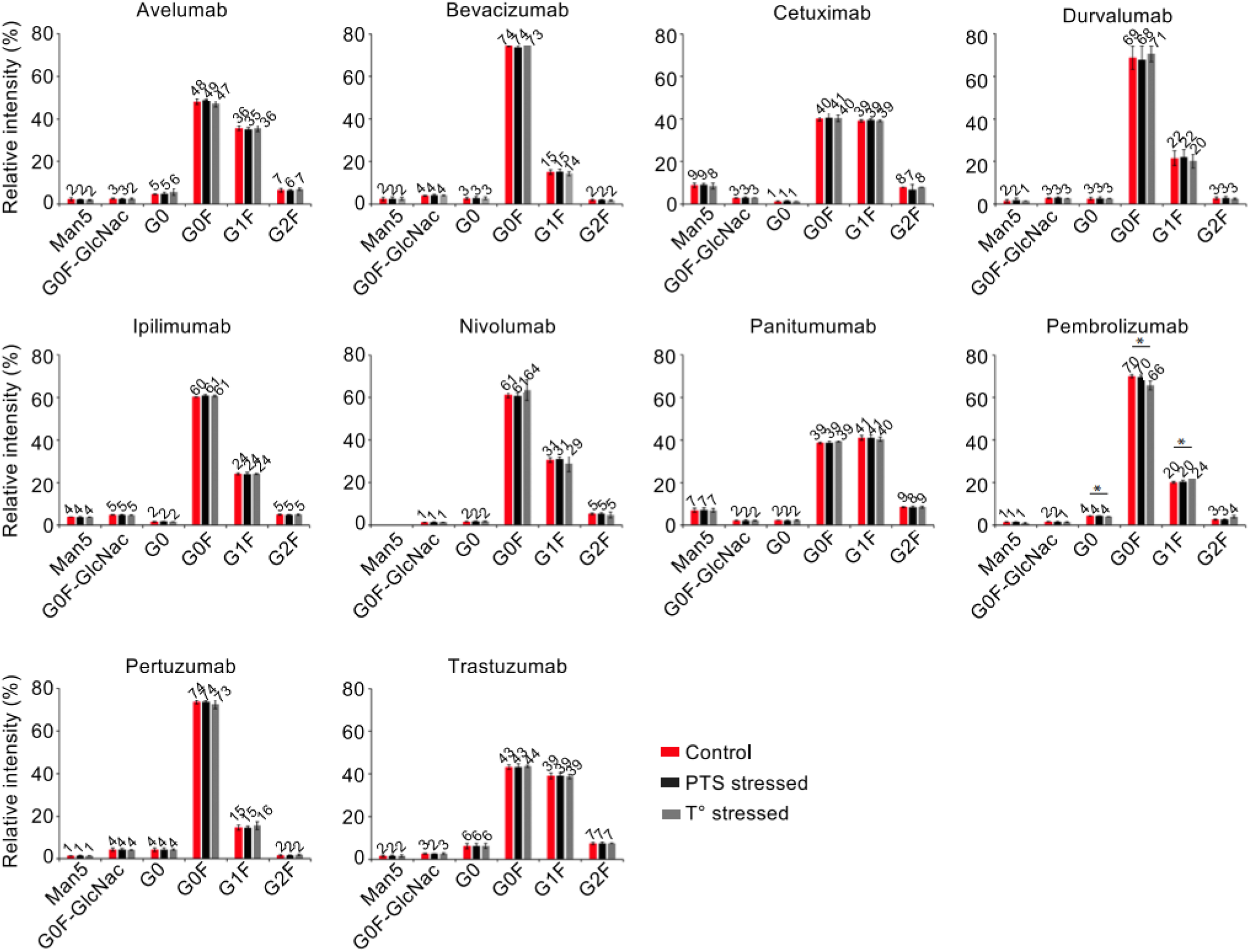
Glycoprofiles of the 11 mAbs after IdeS digestion and rpLC-MS analysis. Relative quantification of the glycoforms was performed after zero-charge deconvolution of MS peaks of Fc/2 subunits. *<u>Red :</u> Reference mAbs* ; *<u>Black :</u> 1 pass-PTS mAbs, <u>Grey :</u> Thermally stressed mAbs*.

### K-clipping

The presence/absence of C-terminal lysine (K-clipping) in the heavy chain (Fc fragment) is another common CQA that reflects the manufacturing consistency. This modification can be assessed by rpLC-MS at the middle level as it generates a mass shift of ∼128 Da (K clipping/deletion variant) and a more hydrophobic variant. Among the 11 mAbs studied, rpLC-MS analysis of the Fc subunit revealed that all of them are formulated in their fully K-clipped versions (>95%, total Fc-K species, **Table 3**), while proteoforms containing the C-terminal lysine representing <5% in diluted mAb samples (**Table 3**). Cetuximab and avelumab are examples of mAbs exhibiting mixtures of K-clipped (∼75%) and non-K-clipped (∼25%) forms. Importantly, the ratio of K-clipped/K-containing species remains unchanged after 1 PTS passage (**Table 3**). Similar conclusions can be drawn from thermally stressed samples, suggesting that deletion of C-terminal lysine is stable once generated from the production process and not significantly impacted upon any of the stress conditions.

**Table 1:** Summary of antibodies tested. We tested a total of 11 monoclonal antibodies (mAbs) using three different batches (except for ipilimumab, which was tested twice with the same batch). The analysis was conducted prior to the expiry date of each mAb.

| Name | Commercial Name | Type | Batches |  |  |  |  |  |
| --- | --- | --- | --- | --- | --- | --- | --- | --- |
|  |  |  | 1st Batch (Exp) |  | 2<sup>nd</sup> Batch |  | 3<sup>rd</sup> Batch |  |
| Atezolizumab | TECENTRIQ® | IgG1 | 1162031 | 08/21 | B0014 | 11/22 | B0016H22 | 03/23 |
| Avelumab | BAVENCIO® | IgG1 | 213079/06 | 03/21 | AU029984 | 12/21 | AU030487 | 03/22 |
| Bevacizumab | MVASIS® | IgG1* | N7407H03 | 12/21 | 1120891 | 03/22 | 1120893 | 03/22 |
| Cetuximab | ERBITUX® | IgG1 | GOOEAJ | 10/23 | GOOH62 | 12/23 | GOOMS1 | 03/24 |
| Durvalumab | IMFINZI® | IgG1 | 0121B16A | 05/21 | 022M17B | 03/23 | 019M17A | 03/23 |
| Ipilimumab | YERVOY® | IgG1 | ABN2184 | 05/22 | ABE3010 | 11/21 | ABE3010 | 11/21 |
| Nivolumab | OPDIVO® | IgG4 | ABQ8316 | 06/21 | ABN601 | 02/22 | CLI10519 | 04/21 |
| Panitumumab | VECTIBIX® | IgG2 | 1113353 | 06/22 | 1115899 | 06/22 | 1121578A | 10/22 |
| Pembrolizumab | KEYTRUDA® | IgG4 | 011736 | 11/21 | T028333 | 01/22 | T031876 | 02/22 |
| Pertuzumab | PERJETA® | IgG1 | H0385H02 | 10/21 | H0437H02 | 07/22 | H0439H05 | 07/22 |
| Trastuzumab | TRAZIMERA® | IgG1 | DE8014 | 02/20 | N3021H02 | 09/22 | DC5104 | 07/23 |

**Table 2:** Orthogonal analysis for Critical Quality Attributes of Monoclonal Antibodies. This table summarizes the critical quality attributes (CQAs) of monoclonal antibodies (mAbs) and the methods used to assess them. SEC n-MS was used to assess aggregation/degradation and to identify post-translational modifications (PTMs), while RPLC-MS was used to detect glycovariants and analyze deamidation, oxidation, and disulfide bonds. Turbidity was assessed using UV/visible spectrophotometry to measure absorbance, and subvisible particles were analyzed using dynamic light scattering to determine the population of particles less than 1 μm in size

| Critical Quality Attributes | Methods | Assessment |
| --- | --- | --- |
| Aggregation/degradation | SEC n-MS | HMWS/LMWS and monomeric pics on Intact mAbs or middle-up analysis |
| Identification of PTM modification | SEC-nMS<br>RPLC-MS | Glycovariants<br>Deamidation, oxydation, disulfide bond analysis |
| Turbidity | UV/visible spectrophotometer | Absorbance |
| Subvisible particules | Dynamic Light Scattering | <1µm particles population |

**Table 3:**
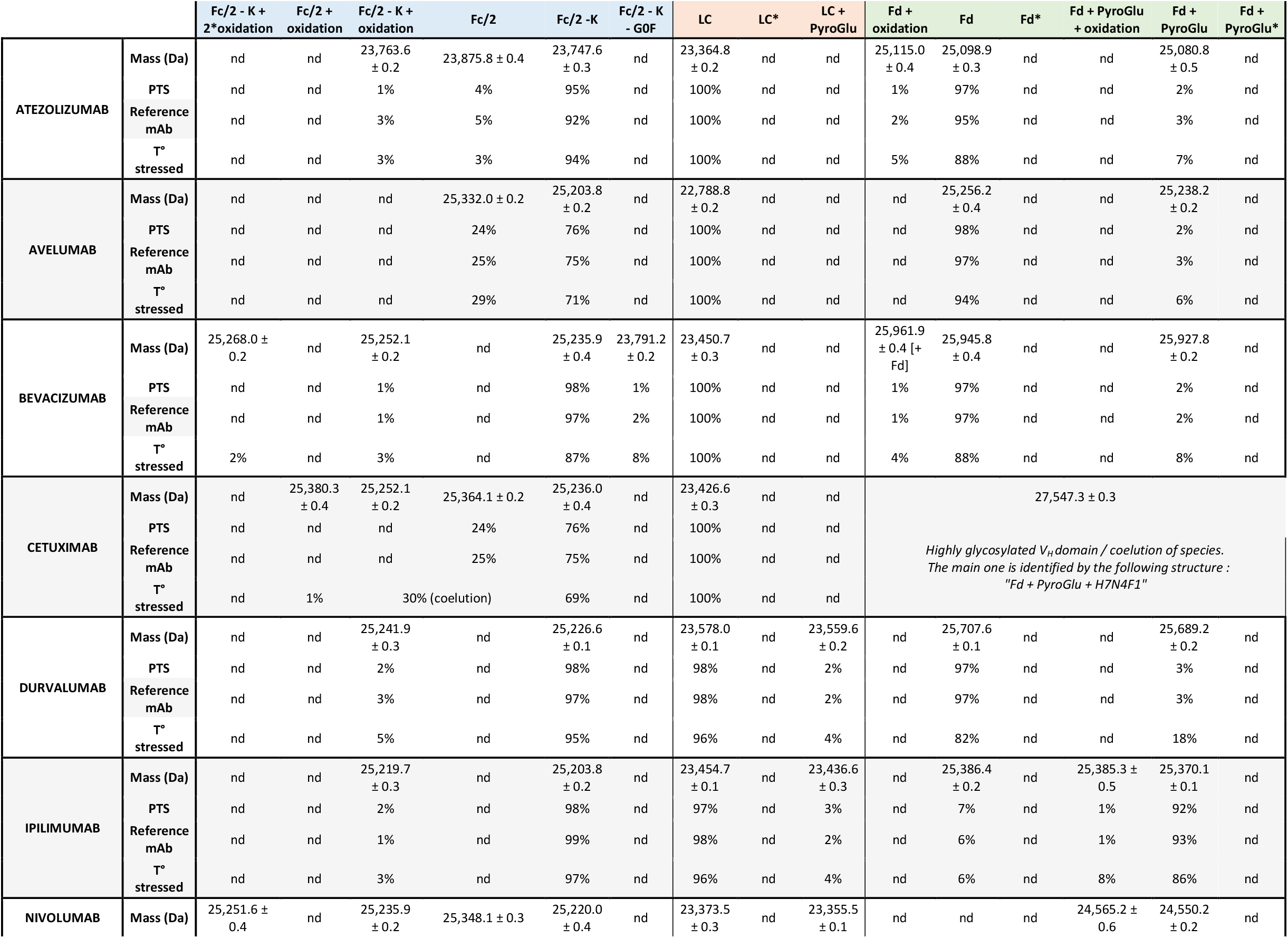
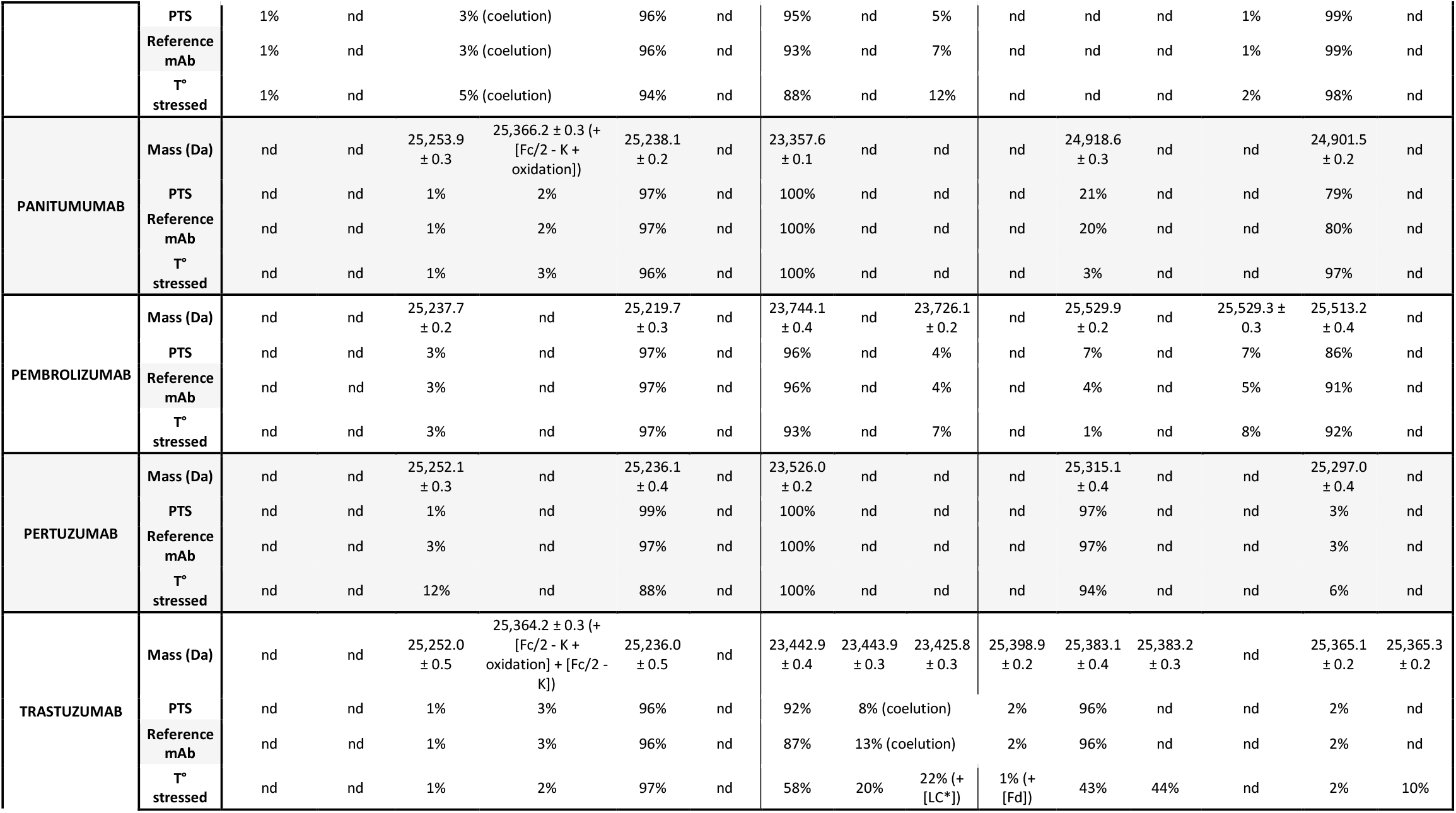
Summary of the middle level rpLC-MS analysis on 11 mAbs.

### Oxidation

Oxidation is an important CQA that impacts the structural integrity, conformational stability, safety, and efficacy of the mAb product^**25**^. While no negative impacts have been reported for the oxidation present on complementary determining regions (CDR) on antigen binding, many negative impacts have been reported for the oxidation of Met residues in the Fc part of the mAb such as decreased thermal stability, increased aggregation, decreased complement dependent cytotoxicity (CDC), decreased binding affinity to neonatal Fc receptor (FcRn), and shorter in vivo half-life, *etc*. We thus sought to explore to which extend PTS passages could induce mAb oxidation (+16 or 32 Da mass increases), knowing that limited effects are expected as air was removed from the plastic packaging. No oxidation was detected on the Fc or Fd subunits of the reference mAbs, at the exception of atezolizumab and bevacizumab for which traces were observed (<3%, **Table 3**). Importantly, these levels were not affected post PTS passage, but were observed for the thermally stressed mAbs (*e.g*. atezolizumab Fd, bevacizumab Fc+Fd, **Table 3**).

### Pyro-Glu

N-terminal pyroglutamate (pyroGlu) is a common mAb modification that mainly results from a non-enzymatic cyclization of N-terminal glutamine (Gln) or to a much lower extent from the conversion of N-terminal glutamate (Glu) into pyroGlu^**26**^. Cyclization of N-terminal Gln/Glu to pyroGlu reduces the molecular weight of a mAb by 17 Da or 18 Da, respectively and induces more acidic variants compared to original non-N-terminally modified mAbs^**20**^. However, the presence of N-terminal pyroGlu exhibits no direct impact on mAb structure and function. As levels of N-terminal pyroGlu observed in mAbs vary strongly with various environmental factors (buffer composition, pH, temperature during cell culture and purification, etc.), we next investigated the N-terminal cyclization on the Fd subunit of the 11 mAbs (**Table 3**). Half of them were formulated as non-pyroGlu forms and very low levels (<3%) of pyroGlu modification on the N-terminal part of our mAbs were detected. At the opposite, the other mAbs are formulated as pyroglutamylated forms (>93%, **Table 3**), except for panitumumab formulated as a mixture of unmodified (21%) and N-terminal pyroGlu (79%) forms. No significant changes were detected after PTS passage, while increased level of pyroGlu forms are systematically observed upon thermal stress (**Table 3**).

### Visible and subvisible particles assessment

Turbidity was investigated by measuring the absorbance at 3 different wavelengths. Comparable relative absorption variations were observed between all conditions which was consistent with the lack of visible foam in the bags as determined by visual inspection (**Table 4**). Subvisible particle formation was then analyzed by dynamic light scattering (DLS) measurements with the same stress conditions applied to the mAbs (control in glass vial, control in plastic bag, thermal stress, and PTS). For the 4 mAbs studied (pertuzumab, pembrolizumab, trastuzumab, and cetuximab), we observed small (5-20 nm) and large (1-10 μm) particles in all conditions tested (**Figure 6**). Our observations indicate that thermally stressed conditions led to a higher abundance of large particles, while no significant differences were observed between the control groups and the PTS conditions. We also observed a third population of particles, ranging from 100-200 nm for each mAb, exclusively in the thermally stressed group.

**Table 4:** Summary of turbidity analysis on 10 mAbs. Relative absorbance (Mean +/-sd) at 550nm, 410nm, and 340nm tested under different conditions. Control represents mAbs in their native state, Heat represents mAbs subjected to thermal stress, and Pneumatic represents mAbs subjected to pneumatic stress. The relative absorbance was measured using a UV/visible spectrophotometer at each wavelength.

| Relative absorbance (Mean +/- sd) | 550nm |  |  | 410nm |  |  | 340nm |  |  |
| --- | --- | --- | --- | --- | --- | --- | --- | --- | --- |
|  | Control | Heat | Pneumatic | Control | Heat | Pneumatic | Control | Heat | Pneumatic |
| TRASTUZUMAB | 0 (+/-0) | 0 (+/-0) | 0 (+/-0) | 0 (+/-0) | 0 (+/-0) | 0,02 (+/-0,03) | 0 (+/-0) | 0 (+/-0) | 0,05 (+/-0,07) |
| ATEZOLIZUMAB | 0,01 (+/-0,02) | 0,07 (+/-0,1) | 0 (+/-0) | 0 (+/-0) | 0 (+/-0) | 0 (+/-0) | 0,08 (+/-0,11) | 0 (+/-0) | 0 (+/-0) |
| AVELUMAB | 0 (+/-0) | 0 (+/-0) | 0 (+/-0) | 0 (+/-0) | 0,16 (+/-0,23) | 0 (+/-0) | 0 (+/-0) | 0 (+/-0) | 0,01 (+/-0,01) |
| BEVACIZUMAB | 0,01 (+/-0,01) | 0 (+/-0) | 0,01 (+/-0,01) | 0,01 (+/-0,01) | 0 (+/-0) | 0,02 (+/-0,01) | 0,02 (+/-0,03) | 0 (+/-0) | 0,05 (+/-0,04) |
| CETUXIMAB | 0 (+/-0) | 0 (+/-0) | 0,28 (+/-0,39) | 0 (+/-0) | 0 (+/-0) | 0,03 (+/-0,05) | 0 (+/-0) | 0 (+/-0) | 0,46 (+/-0,65) |
| DURVALUMAB | 0 (+/-0) | 0 (+/-0) | 0 (+/-0) | 0 (+/-0) | 0 (+/-0) | 0 (+/-0) | 0,06 (+/-0,08) | 0 (+/-0) | 0 (+/-0) |
| NIVOLUMAB | 0 (+/-0) | 0 (+/-0) | 0 (+/-0) | 0 (+/-0) | 0 (+/-0) | 0 (+/-0) | 0 (+/-0) | 0 (+/-0) | 0 (+/-0) |
| PANITUMUMAB | 0,01 (+/-0,02) | 0 (+/-0) | 0,01 (+/-0,01) | 0 (+/-0) | 0 (+/-0) | 0,01 (+/-0,01) | 0,05 (+/-0,06) | 0 (+/-0) | 0,01 (+/-0,01) |
| PEMBROLIZUMAB | 0,01 (+/-0,01) | 0 (+/-0) | 0,01 (+/-0,01) | 0,01 (+/-0,01) | 0,76 (+/-1,07) | 0,01 (+/-0,02) | 0,01 (+/-0,01) | 0 (+/-0) | 0,02 (+/-0,02) |
| PERTUZUMAB | 0 (+/-0) | 0,05 (+/-0,08) | 0 (+/-0) | 0 (+/-0) | 0,07 (+/-0,1) | 0 (+/-0) | 0,03 (+/-0,04) | 0,12 (+/-0,17) | 0 (+/-0) |

**Figure 6:**
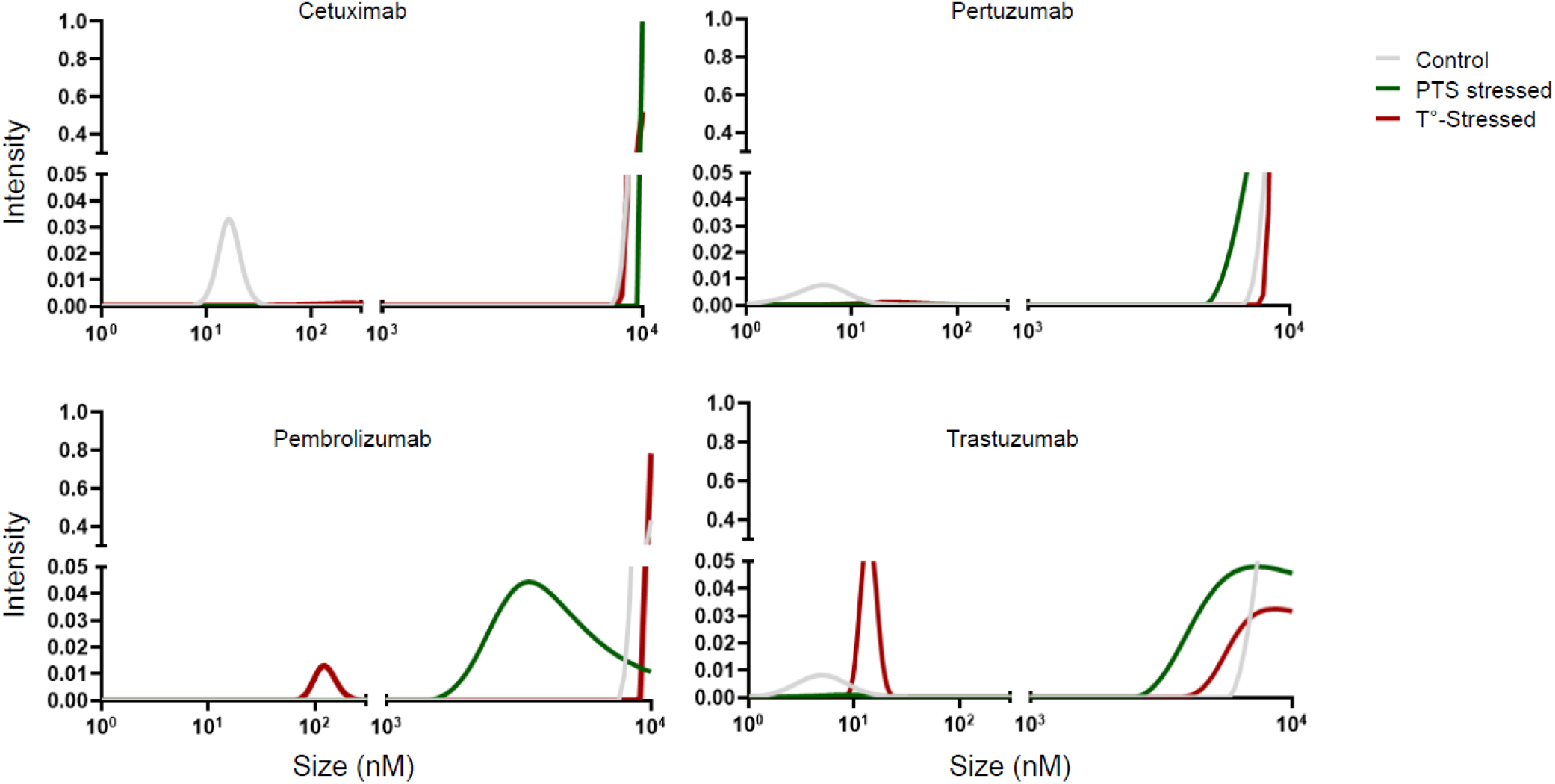
Dynamic Light Scattering analysis of four stressed mAbs. Two distinguishable particle populations were observed: Small particles ranging from 5-20nm and larger particles ranging from 1-10μm. A third population emerged under thermally stressed conditions, consisting of particles ranging from 100-200nm for each monoclonal antibody.

## Discussion and conclusion

Quality issues due to biologics were highlighted by the development of trastuzumab biosimilars where batch-to-batch differences of the reference product were monitored over a long-term period of time to assess the ranges of variations for key CQAs^27–29^. A modification of the glycosylation profile was observed; such modifications impacted the FcγRIIIa binding as well as the ADCC activity of the mAbs without any decreasing their antiproliferative activity against the HER2-positive cancer cells^18,30^. However, in a phase III clinical trials aiming to validate the equivalence between the referent trastuzumab and SB3^31^ or ABP980^32^, the predefined equivalence margins to assess the equivalence of the primary efficacy endpoints (pathological complete response – pCR) were exceeded. The results of this clinical trial suggested that a potential superiority could not be ruled out. A deeper analysis of the results of the clinical trial demonstrated that the pCR rates were dictated by an unexpected modification of the trastuzumab reference product for which alterations of fucose and mannose levels were determined.

This prompted us to focus on some major CQA to assess the impact of PTS passage in our institute with the final objective to develop analytical methods that could be utilized in hospitals equipped with qualified LC-MS systems by focusing on methods without extensive mAb handling as well as with protocols that could be automatized. Intact protein analysis with SEC-nMS was selected as first line method to evaluate multiple quality attributes within a single 10 min run. The SEC-nMS method allows assessment of the drug product purity with simultaneous detection, quantification, and accurate mass identification of the main product (including glycoprofile monitoring) along with aggregates (HMWS) and fragments (LMWS) identification. For more precise analysis of CQA related to important PTMs (glycosylation, oxidation, pyroglytamylation, and C-terminal K cleavage), complementary rpLC-MS analyses of IdES downsized mAb subunits were performed. Altogether, the intact- and middle-level analytical pipelines demonstrated that PTS did not induced major changes both in terms of size variants and for a selected series of charge variants.

This work suggests that compared to bottom up workflows (peptide mapping), which are more difficult to envision in a QC laboratory of a hospital, intact- and middle-level LC-MS analysis could be proposed as “multiple attribute monitoring” methods in regulatory environments like hospital for routine analysis. The external stress induced by PTS on a mAb in its diluted version potentially represents a more sensitive condition gathering high risks regarding the preservation of the integrity of the drug product. Hence, the orthogonal analysis of the 11 mAbs diluted in a version ready to be injected to patients demonstrated the absence of observed impact induced by the PTS as analyzed with the major CQA methods (HMWS, LMWS, glycoform profile, oxidation, etc). A comparable aggregation/degradation status and absence of alterations addressing the protein primary structure, the glycosylation profile, the oxidation level, N-terminal cyclization and C-terminal K cleavage along with identical particle sizes were observed in the PTS.

A key confounding factor related to the risk assessment is whether air is removed from the plastic bag. Indeed, here, at the opposite to what was previously performed^6^, the air was removed from the plastic bag to reduce as much as possible the air-liquid interface to minimize the risks of oxidations. This modification in the preparation method before using the PTS seems to be the most important factor to justify using the PTS to transport the mAbs in a routine environment. Hence, through the evaluation of the 11 mAbs, the PTS did not modify any of the mAbs. However, caution should be considered to extend these results to PTS at every hospital without previous assessment of the configuration of their transportation system. If the PTS configuration highly differs from ours, a case-by-case quality control of the PTS should be recommended. Nevertheless, the results obtained with 1 passage and 10 passages providing comparable findings, it is safe to assume that PTS should not induce modification of the mAbs in respect to the CQA if the air-liquid interface is reduced at the minimum level.

## Material and methods

### Materials

A total of 11 mAbs were obtained from the commercial final products provided by the ICANS hospital pharmacy. Three different batches of each one, with the 3 defined conditions, were used for the analysis following orthogonal methods for different critical quality attributes (CQAs) (**Table 1** and **2**).

### Reference mAbs

Commercially non-diluted mAbs stored in their original glass packaging were used for analysis with 3 different batches.

### Pneumatic stress condition

Each batch was diluted in a prefilled sodium chloride polyolefin (PO) bag at a concentration of 1 mg/ml. The air was removed after dilution limiting the impact of the air-water interface. The bags were then placed in the pneumatic cartridge and underwent one trip through the PTS (Aerocom AC4000; Krautergersheim, France) before analysis.

### Thermal stress condition

Each batch, at commercial concentration, was incubated at 50°C during 20 days in the original glass vial before analysis to generate the thermal stress.

### Acceleration measurements

As a qualification process for the PTS, we have established a cartography of stress constraints induced during the transport. An accelerometer (Volcraft^®^ DL-131g, Hirschau, Germany) was fixed into a plastic bag in the cartridge used over a PTS transportation. A total of 10 travels were performed providing a characterization of all forces induced by the PTS on the bag. The average speed of the cartridge was fixed at 3.5m/s.

### SEC-nMS

An ACQUITY UPLC H-class system (Waters, Manchester, UK) comprising a quaternary solvent manager, a sample manager cooled at 10 °C, a column oven maintained at room temperature and an UV detector operating at 280 nm and 214 nm hyphenated to a Synapt G2 HDMS mass spectrometer (Waters, Manchester, UK) was used for the online SEC-native MS instrumentation. Forty micrograms of mAb were loaded on the ACQUITY UPLC Protein BEH SEC column (4.6 × 150 mm, 1.7 μm particle size, 200 Å pore size) from Waters (Manchester, UK) using an isocratic elution of 100 mM ammonium acetate (NH_4_OAc) at pH 6.9 with the following flow rate gradiant : 0.250 mL/min over 3.0 minutes, then 0.100 mL/min from 3.1 to 15.0 minutes, and finally 0.250 mL/min from 15.1 to 18.0 minutes.

The Synapt G2 HDMS was operated in positive mode with a capillary voltage of 3.0 kV while sample cone and pressure in the interface region were set to 180 V and 6 mbar, respectively. Acquisitions were performed in 50–15,000 m/z range with a 1.5 s scan time. The mass spectrometer was calibrated using singly charged ions produced by a 2 g/L solution of cesium iodide (Acros organics, Thermo Fisher Scientific, Waltham, MA USA) in 2-propanol/water (50/50 v/v). Native MS data interpretations were performed using Mass Lynx V4.1 (Waters, Manchester, UK).

Relative quantification of HMWS/LMWS by intact SEC-native was performed using the relative abundances calculated from the areas of the chromatographic peaks obtained at 280 nm. For the quantification of glycoforms at intact levels, the relative abundances were based on the intensities obtained after deconvolution of the mass spectrum. Standard deviations were calculated using biological replicates.

### Middle level analysis using rpLC-MS

Enzymatic digestion (FabRICATOR®, Genovis, Sweden) was performed by incubating one unit of IdeS protease per microgram of mAb. First of all, the enzyme was reconstituted at four units per microliter in ultrapure water. Then, the corresponding volume was added to fifty micrograms of mAb. The total volume was completed at seventy microliters by adding a buffer composed of 50 mM Na_2_HPO_4_ and 150 mM NaCl at pH 6.7. Finally, the mixture was incubated at 37 °C for thirty minutes under shaking (500 rpm). In order to perform a strong denaturation, thirty-three milligrams of guanidine HCl (powder) was directly added to the mixture. The reduction step was performed by adding ten microliter of TCEP (tris(2-carboxyethyl)phosphine) solution at 560 mM. After sixty minutes incubation at 57 °C under shaking (500 rpm), the reaction was quenched by adding one microliter of TFA (trifluoroacetic acid) solution. LC-MS analyses were performed using an Agilent 1200 series coupled to a maXis II (Bruker) Q-TOF based mass spectrometer. A volume equivalent to five micrograms of sample preparation was injected on a BioResolve™ RP mAb 2.1 mm internal diameter and 150 mm length, pore size 450 Å and particle size 2.7 μm, polyphenyl column (Waters, Manchester, UK) set at 80 °C. The UV chromatogram was acquired with a DAD detector operating from 200 to 400 nm. Ultrapure water for mobile phase A (0.10% TFA) and acetonitrile for mobile phase B (0.08% TFA) were used to perform the chromatographic separation. The gradient was generated at a flow rate of 300 μL/min and the elution program started at 25% mobile phase B during two minutes to reach 44% in thirty-eight minutes. Then, five minutes of washing step at 75% and finally fourteen minutes of equilibration at 25% mobile phase B. A blank was injected between each sample under the same conditions. The mass spectrometer was operated in positive mode with a capillary voltage of 4,500 V and CID value was set at 70 eV. Acquisitions were performed on the mass range 500-3,000 m/z. Calibration was performed using the singly charged ions produced by a solution of 2 g/L caesium iodide in 2-propanol/water (50/50 v/v). Data interpretation was performed by using Compass DataAnalysis 4.3 software (Bruker Daltonics). Since the molecular isotopic cluster was not fully resolved, average molecular masses were measured.

Relative quantification of PTMs identified from middle level rpLC-MS analysis was performed using the relative abundances calculated from the areas of the chromatographic peaks obtained at 280 nm. For the quantification of glycoforms at the middle level, the relative abundances were based on the intensities obtained after deconvolution of the mass spectrum. Standard deviations were calculated using biological replicates.

### Visible particles

Absorbance was measured with a spectrophotometer (UV-1800, Shimadzu) at 340, 410 and 550nm for each condition (control diluted in PO bag, pneumatic stress condition with one travel through the PTS, and thermal stress condition). Absorbance measurements were performed by aliquoting 1 ml of each sample and were expressed relative to the blank condition (sodium chloride in PO bag). A visual inspection of each bag transported with the PTS system was also implemented to detect foam.

### Subvisible particles

The mAbs were stored under 3 different conditions (control in vials, pneumatic stress condition with one trip through the PTS, and thermal stress condition) and then the effect of these conditions on their size was evaluated by analyzing dynamic light scattering (DLS) measurements (Amerigo Particle Size Analyzer, Cordouan Technologies). The DLS measurements were performed by directly aliquoting the samples into a cuvette exposed to an external laser of 638 nm. The raw data were further analyzed by using the sparse Bayesian learning (SBL) algorithm and then the intensity average number of particles was plotted on Graphpad Prism 8.0.

### Statistical analysis

To evaluate significant differences between the reference monoclonal antibody (mAb) condition and the conditions of thermal or pressure thermal shock (PTS), an analysis of variance (ANOVA) test was conducted. To further examine the differences between the groups, the ANOVA test was supplemented with a Tukey test for intergroup comparisons. Differences were considered statistically significant if the p-value was less than 0.05

## Data Availability

All data produced in the present study are available upon reasonable request to the corresponding author.

## Acknowledgments

This study was supported by the CNRS, the University of Strasbourg, the Agence Nationale de la Recherche, the French Proteomic Infrastructure (ProFI; ANR-10-INBS-08-03), the Interdisciplinary Thematic Institute IMS (Institut du Médicament Strasbourg), as part of the ITI 2021-2028 supported by IdEx Unistra (ANR-10-IDEX-0002), SFRI-STRAT’US project (ANR-20-SFRI-0012), and by the Institut de Cancérologie Strasbourg Europe.

Mass spectrometers were purchased through financial support of GIS IBiSA, Région Grand Est, the IdeX initiative of the University of Strasbourg and the French National Proteomic Infrastructure.

Authors would like to acknowledge the fruitful discussion with Aerocom^ltd^ (Krautergersheim, France) about their PTS system which led to this research project.

## Author contributions

Pierre Coliat have full access to all the data in the study and take responsibility for the integrity of the data and accuracy of the data analysis.

Concept and design: Pierre Coliat, Martin Demarchi, Xavier Pivot, Alexandre Detappe, Sarah Cianférani

Supervision: Pierre Coliat, Sarah Cianférani, Alexandre Detappe

Statistical analysis: Pierre Coliat, Sarah Cianférani

Administrative, technical, or material support: Dan Karouby, Stéphane Erb, Hélène Diemer

Acquisition, analysis, and interpretation of data: Pierre Coliat, Alexandre Detappe, Stéphane Erb, Hélène Diemer, Mainak Banerjee, Chen Zhu

Manuscript draft: Pierre Coliat, Sarah Cianférani

Critical revision of the manuscript for important intellectual content and approval: Pierre Coliat, Alexandre Detappe, Dan Karouby, Stéphane Erb, Hélène Diemer, Mainak Banerjee, Chen Zhu, Martin Demarchi, Sarah Cianférani, Xavier Pivot

