## Supplemental Figure 1 for "Clinical validation of pneumatic transportation systems for monoclonal antibodies"

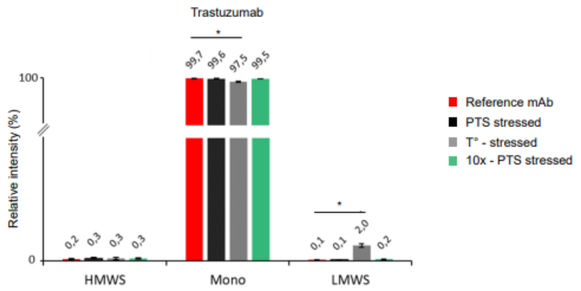


**Supplemental Figure 1 : Quantification of HMWS and LMWS from intact SEC-native MS analysis on trastuzumab**. Histograms represent SEC-UV peak area integration of aggregates (HMWS, mostly dimers) and fragments (mostly Fab-FC and Fab) for trastuzumab. *Red : Reference mAbs ; Black : 1 pass-PTS mAbs, Grey : Thermally stressed mAbs; Green: 10x pass-PTS mAbs.*
